# Impact of the establishment of a national lead agency on the emergency medical service system in Khon Kaen Province, Thailand

**DOI:** 10.1101/2025.02.13.25321942

**Authors:** Papawadee Yapanan, Tawatchai Impool, Ratrawee Pattanarattanamolee, Wuttikul Thanakanjanaphakdee, Shinichi Tokuno, Shinji Nakahara

## Abstract

**Purpose:** National Institute for Emergency Medicine (NIEM) was established as the national lead agency for emergency medicine in 2008 in Thailand. This study aimed to evaluate the longitudinal effect of establishing NIEM on emergency medical service (EMS) use and case fatalities among patients with severe trauma.

**Method:** Data were extracted from Khon Kaen Hospital’s Injury Surveillance Database. The analysis included injured patients with an Injury Severity Score of >15, directly transferred to its emergency department between January 1997 and December 2021. Changes in formal EMS use, total EMS use, and in-hospital deaths were investigated using segmented regression analysis. The change point in 2008 divided the study period into two segments.

**Result:** The proportion of formal EMS use among severely injured patients increased by approximately 1.5% annually before 2008, with a level change of 52.9 percentage point (*pp*) increases in 2008, and without a significant slope change. The total EMS use, including informal EMS, increased by approximately 3.4% annually before 2008, with a level change of 17.9 *pp* increase in 2008, and a significant slope change in the decreasing direction of the increasing trend. The proportion of in-hospital deaths showed no significant changes before 2008, with an increase of 8.0 *pp* in 2008, with significant slope change and decreasing trends after 2008.

**Conclusions:** The trends indicated a significant increase in EMS use among patients with severe trauma after establishing NIEM in 2008, which reflects the integration of informal EMS units into the formal system. However, a reduction in fatalities in 2008 was not observed.

## Introduction

Emergency care is a crucial component of healthcare system. Prioritizing a comprehensive approach to early recognition and resuscitation of critically unwell patients significantly reduces morbidity and mortality.(1) Emergency care includes emergency medical services (EMS) and in-hospital treatments. The development of emergency care systems in low- and middle-income countries is often limited by resources.(2, 3) In addition, lack of accessibility and availability of emergency care services is associated with higher morbidity and further deterioration of health, leading to higher long-term care costs.(4) However, improving emergency care systems can reduce preventable mortality and morbidity and is cost-effective, especially in low- and middle-income countries.(5) The World Health Assembly has called on member states to prioritize the establishment of integrated emergency and trauma care systems.(1)

The national lead agency and its leadership are key elements in developing an EMS system that can deliver high-quality care. The lead agency should develop policies, regulations, operational standards (guidelines/protocols), communication and information systems, standardized education curriculum, and accreditation system for human resource development, evaluate the operations to assure care quality, and coordinate with other agencies.(6) In Thailand, National Institute for Emergency Medicine (NIEM) was established under the Emergency Medical Act in 2008.(7, 8)

Although a national lead agency and its leadership are key indicators of a mature EMS system,(9-11) few studies have investigated the impact of establishing a national lead agency for EMS and its policy development in Thailand;(7, 12) no such studies have been conducted in other countries. Suriyawongpaisal et al.(12) described the coverage and response time of the EMS in Thailand after the NIEM establishment and made a simple comparison with pre-establishment data. Pochaisan et al.(7) described trends in the coverage and number of EMS units only graphically over approximately 10 years before and after the establishment of the NIEM. However, these studies did not conduct appropriate time-series analyses to evaluate trend changes using data from sufficiently long periods before and after establishment.

This study aimed to determine the impact of an enhanced EMS system through NIEM establishment on EMS use and case fatalities among patients with severe trauma using data from the Emergency Department of Khon Kaen Hospital in Thailand. We used a segmented regression of interrupted time-series analysis, which is more appropriate than a simple before-after comparison.(13) The results of the present study will be particularly beneficial to low- and middle-income countries developing EMS systems, as establishment of the national lead agency will greatly improve the systems.

## Research methods

### 1. Study design

This retrospective observational study used anonymized data from the Injury Surveillance Database of the Khon Kaen Hospital. Time series analysis was performed to investigate trend changes before and after NIEM establishment from 1997 to 2021 (NIEM was established in 2008) using annually aggregated data on EMS use proportion and in-hospital mortality (case fatality rate). The study protocol was approved by the Ethics Committee of Khon Kaen Hospital and the Graduate School of Health Innovation, Kanagawa University of Human Services.

### 2. Study settings

Khon Kaen Hospital is the leadingregional hospital in the northeast region of Thailand that provides tertiary care and receives referred patients from the district and surrounding provincial-level hospitals. Khon Kaen Hospital has an emergency command control and information center that coordinates and communicates with the EMS units and hospitals in Khon Kaen Province.(14) In 1993, the Accident Trauma Center at Khon Kaen Hospital was established with support from the Japan International Cooperation Agency (JICA); hospital-based EMS units were also established. Since then, Khon Kaen Hospital has led the development of the Thai EMS system as a model province.

The EMS in Thailand was initiated by voluntary foundations, and to date, there are a number of volunteer EMS units.(15) Establishment of hospital-based EMS units has gradually progressed since the 1990s but has not adequately covered the population. Since 2008, volunteer EMS units have been formally trained, registered, and integrated into the EMS system, and basic EMS units have been stationed at sub-district offices. Currently, different levels of EMS units are operated under the coordination of command control centers: the basic units are stationed in communities with basic personnel and equipment, and advanced units are stationed at hospitals with paramedics and physicians.

Khon Kaen Hospital’s Injury Surveillance Database registers all injured patients treated in the hospital’s emergency department. Registered information included time, place, activities, causes, diagnoses, injuries severity, transportation mode to the hospital, physiological indicators at hospital arrival, disposition from the emergency department, and outcomes (e.g., in-hospital death and discharge alive). Diagnoses were categorized using the International Classification for Disease, 10^th^ revision, and severity was categorized using the Abbreviated Injury Scale (AIS) severity score, which ranged from 1 (minor) to 6 (critical).(16)

### 3. Study participants

The time series analysis included adult patients (aged 15–84 years) with an Injury Severity Score (ISS) of >15 who were treated in the emergency department of Khon Kaen Hospital between January 1997 and December 2021. The exclusion criteria were age ≥ 85 years (very old patients, few in number, and having high fatality rates regardless of injury severity), those referred from another hospital (because this study focused on EMS use at the scene), non-traumatic injuries (e.g., poisoning and drowning), those with cardiac arrest at hospital arrival, and those with data lacking in ISS, age, referral, and injury type (Figure 1).In total, 5609 patients were included in the analysis of EMS use. Cardiac arrest cases at hospital arrival were excluded from the analysis for case fatality, which included 5409 patients.

**Figure 1.**
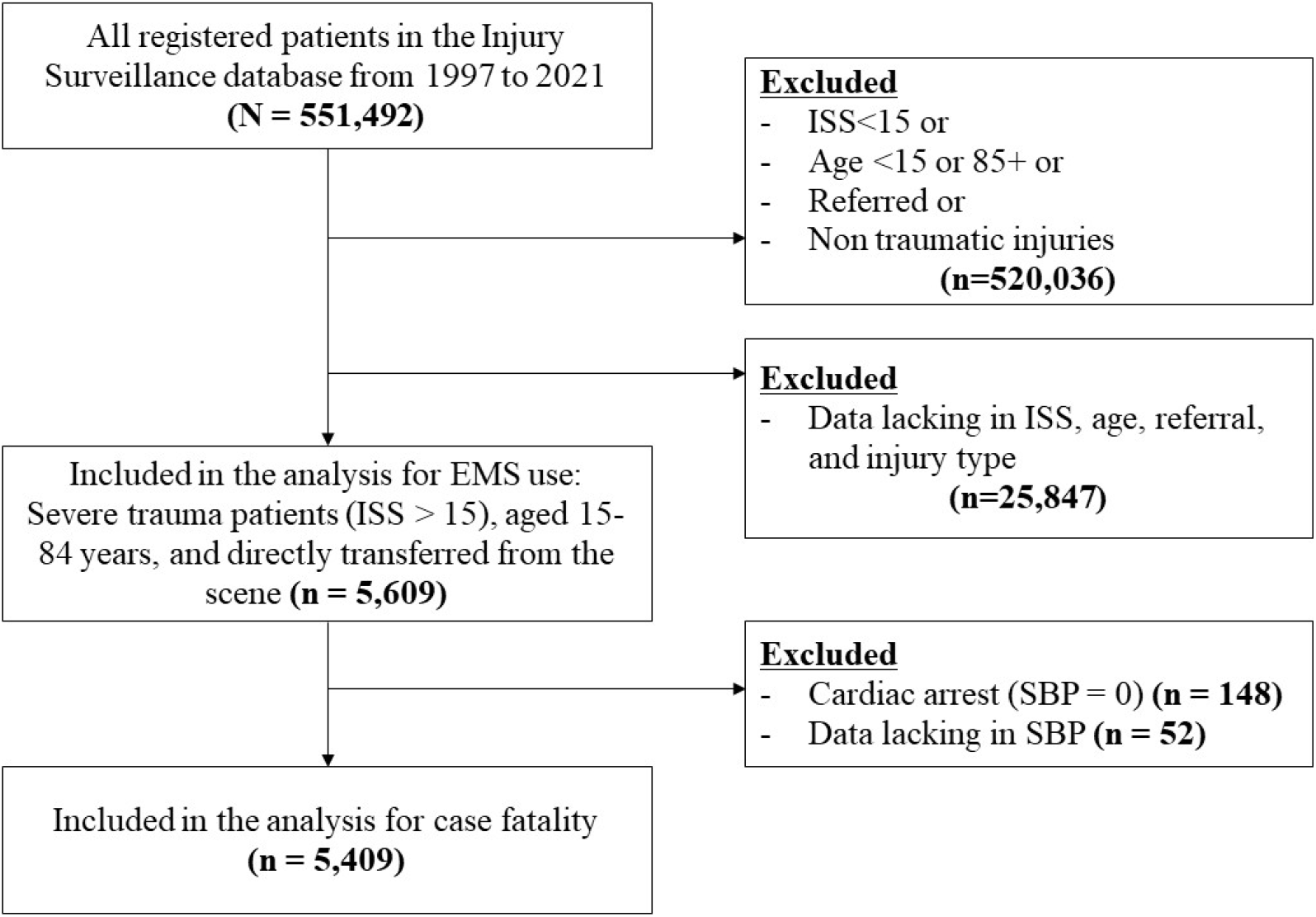
Flow diagram of participant selection

### 4. Variables

The following information was extracted from the injury surveillance data: transportation of the patients (formal EMS, informal EMS, or non-EMS), Age, Sex, AIS severity score, ISS, and status at discharge. Because ISS data were not available between 1997-2007, they were calculated based on the AIS during this period.

The outcome variables for the time series analysis were the proportion of formal EMS use, total EMS use (the numerator included both formal and informal EMS use), and case fatalities (in-hospital deaths divided by the number of patients analyzed). The independent variable of the time-series analysis was the years (1997–2021) with a change point in 2008 (NIEM establishment) that divided the period into two segments.

### 5. Sample size

The sample size for the interrupted time-series analysis was 25 years of data from Khon Kaen Hospital from 1997 to 2021, with a change point in 2008 (NIEM establishment), which divides the period into two segments.

### 6. Statistical analysis

Segmented regression analysis is a powerful statistical method for estimating the intervention effects in interrupted time-series studies.(17) A time series is a sequence of values for a particular measure obtained at regularly spaced intervals over time. Segments in a time series are defined when the sequence of measures is divided into two portions at change points; this is the national lead agency establishment in this study. This analysis is a method for statistically modeling interrupted time-series data to draw more formal conclusions about the impact of an intervention or policy.

The regression model is indicated as follows:

Y_t_ = β_0_ + β_1_t + β_2_x + β_3_xt,

where

Y: proportion at a given time point

β_0_: intercept (constant) β_1_: pre-change slope

β_2_: level change

β_3_: slope change t: time

x: time after change (x=1 after the change point)

(The post-change slope is given by β_1_ + β_3_)

All statistical analyses were performed using Stata version 17 (Stata Corp LLC, Texas, USA). Annual data between 1997-2021 were used to conduct a segmented regression with a change point in 2008 (when the NIEM was established). The ITSA command with Newey–West standard errors and one lag was used for the time-series analysis, and the Cumby-Huizinga test was used for testing autocorrelation. To determine the appropriate lag level, we conducted this analysis with no lag.

## Results

### Patient characteristics

Patients with severe trauma visiting the emergency department of Khon Kaen Hospital increased from around 200 in the 1990s and 2000s to around 300 in the late 2010s (Table 1). Most patients were male (81.4%). The median age tended to increase, and the median ISS decreased during the study period. The percentage of formal EMS use, which involved transportation by registered EMS units with well-trained personnel, increased over time (Table 2). Before 2008, informal EMS units from voluntary charity organizations and the private sector transported many patients to Khon Kaen Hospital. Immediately after the 2008 NIEM establishment, informal EMS rapidly decreased. The percentage of in-hospital deaths unevenly reduced between 1997 and 2021.

**Table 1.**
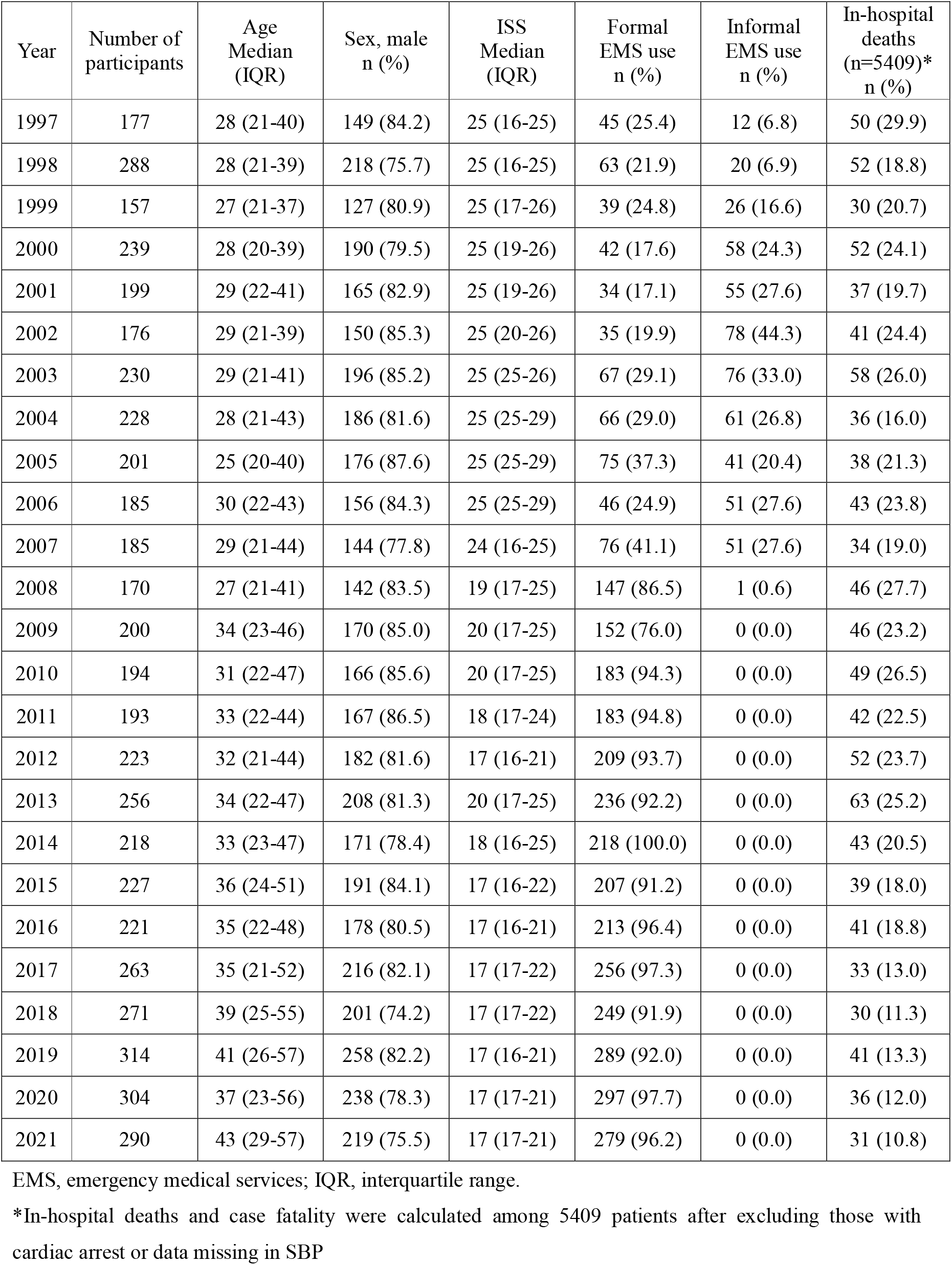
Participant characteristics, EMS use, and in-hospital deaths (n=5,609)

| Year | Number of participants | Age Median (IQR) | Sex, male n (%) | ISS Median (IQR) | Formal EMS use n (%) | Informal EMS use n (%) | In-hospital deaths (n=5409)* n (%) |
| --- | --- | --- | --- | --- | --- | --- | --- |
| 1997 | 177 | 28 (21-40) | 149 (84.2) | 25 (16-25) | 45 (25.4) | 12 (6.8) | 50 (29.9) |
| 1998 | 288 | 28 (21-39) | 218 (75.7) | 25 (16-25) | 63 (21.9) | 20 (6.9) | 52 (18.8) |
| 1999 | 157 | 27 (21-37) | 127 (80.9) | 25 (17-26) | 39 (24.8) | 26 (16.6) | 30 (20.7) |
| 2000 | 239 | 28 (20-39) | 190 (79.5) | 25 (19-26) | 42 (17.6) | 58 (24.3) | 52 (24.1) |
| 2001 | 199 | 29 (22-41) | 165 (82.9) | 25 (19-26) | 34 (17.1) | 55 (27.6) | 37 (19.7) |
| 2002 | 176 | 29 (21-39) | 150 (85.3) | 25 (20-26) | 35 (19.9) | 78 (44.3) | 41 (24.4) |
| 2003 | 230 | 29 (21-41) | 196 (85.2) | 25 (25-26) | 67 (29.1) | 76 (33.0) | 58 (26.0) |
| 2004 | 228 | 28 (21-43) | 186 (81.6) | 25 (25-29) | 66 (29.0) | 61 (26.8) | 36 (16.0) |
| 2005 | 201 | 25 (20-40) | 176 (87.6) | 25 (25-29) | 75 (37.3) | 41 (20.4) | 38 (21.3) |
| 2006 | 185 | 30 (22-43) | 156 (84.3) | 25 (25-29) | 46 (24.9) | 51 (27.6) | 43 (23.8) |
| 2007 | 185 | 29 (21-44) | 144 (77.8) | 24 (16-25) | 76 (41.1) | 51 (27.6) | 34 (19.0) |
| 2008 | 170 | 27 (21-41) | 142 (83.5) | 19 (17-25) | 147 (86.5) | 1 (0.6) | 46 (27.7) |
| 2009 | 200 | 34 (23-46) | 170 (85.0) | 20 (17-25) | 152 (76.0) | 0 (0.0) | 46 (23.2) |
| 2010 | 194 | 31 (22-47) | 166 (85.6) | 20 (17-25) | 183 (94.3) | 0 (0.0) | 49 (26.5) |
| 2011 | 193 | 33 (22-44) | 167 (86.5) | 18 (17-24) | 183 (94.8) | 0 (0.0) | 42 (22.5) |
| 2012 | 223 | 32 (21-44) | 182 (81.6) | 17 (16-21) | 209 (93.7) | 0 (0.0) | 52 (23.7) |
| 2013 | 256 | 34 (22-47) | 208 (81.3) | 20 (17-25) | 236 (92.2) | 0 (0.0) | 63 (25.2) |
| 2014 | 218 | 33 (23-47) | 171 (78.4) | 18 (16-25) | 218 (100.0) | 0 (0.0) | 43 (20.5) |
| 2015 | 227 | 36 (24-51) | 191 (84.1) | 17 (16-22) | 207 (91.2) | 0 (0.0) | 39 (18.0) |
| 2016 | 221 | 35 (22-48) | 178 (80.5) | 17 (16-21) | 213 (96.4) | 0 (0.0) | 41 (18.8) |
| 2017 | 263 | 35 (21-52) | 216 (82.1) | 17 (17-22) | 256 (97.3) | 0 (0.0) | 33 (13.0) |
| 2018 | 271 | 39 (25-55) | 201 (74.2) | 17 (17-22) | 249 (91.9) | 0 (0.0) | 30 (11.3) |
| 2019 | 314 | 41 (26-57) | 258 (82.2) | 17 (16-21) | 289 (92.0) | 0 (0.0) | 41 (13.3) |
| 2020 | 304 | 37 (23-56) | 238 (78.3) | 17 (17-21) | 297 (97.7) | 0 (0.0) | 36 (12.0) |
| 2021 | 290 | 43 (29-57) | 219 (75.5) | 17 (17-21) | 279 (96.2) | 0 (0.0) | 31 (10.8) |

EMS, emergency medical services; IQR, interquartile range.
\*In-hospital deaths and case fatality were calculated among 5409 patients after excluding those with cardiac arrest or data missing in SBP

**Table 2.**
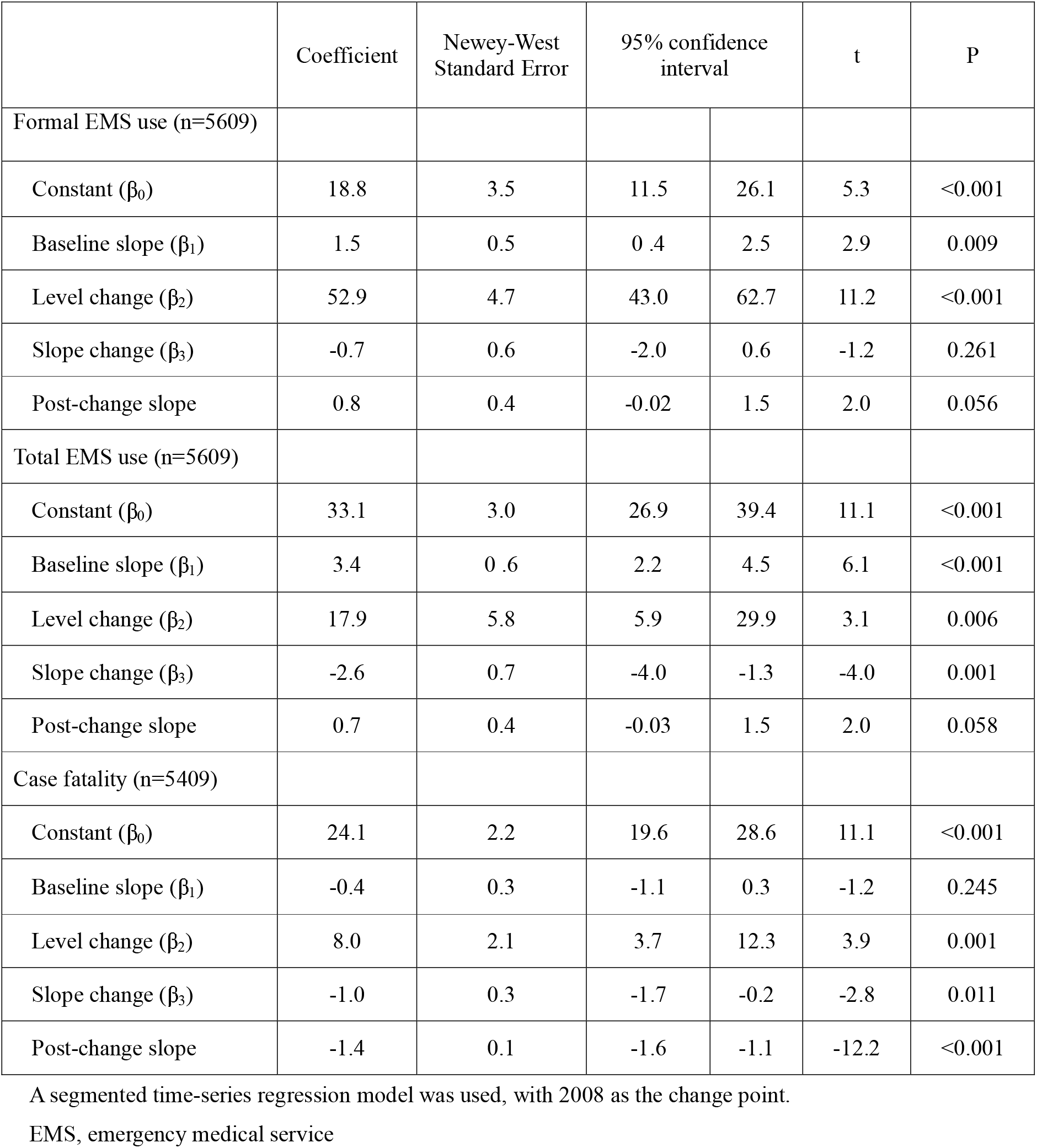
Time series analysis of EMS use and case-fatalities among severe trauma patients 1997-2021.

### Time series analyses

No autocorrelation was detected in the time-series analyses, regardless of the lag level, and the Cumby-Huizinga test for autocorrelation indicated no serial correlations in all analyses. The results of the time-series analyses did not differ with or without lag. However, we presented the results of the analysis with a single lag.

In the analysis of formal EMS usage trends from 1997-2021, in the beginning, the percentage of EMS usage was approximately 20% (Figure 2a and Table 1). Before the NIEM establishment, the percentage increased by approximately 1.5% (95% CI=0.4–2.5) annually (Table 2). The level change in 2008 was significant: a 52.9 percentage point increase (95% CI=43.0–62.7). The estimated slope change in 2008 was small and insignificant. After NIEM establishment, the percentage of formal EMS trend slope was significantly increased 0.8% yearly. By 2021, the percentage of formal EMS usage had increased to 96.2% (Table 1).

**Figure 2.**
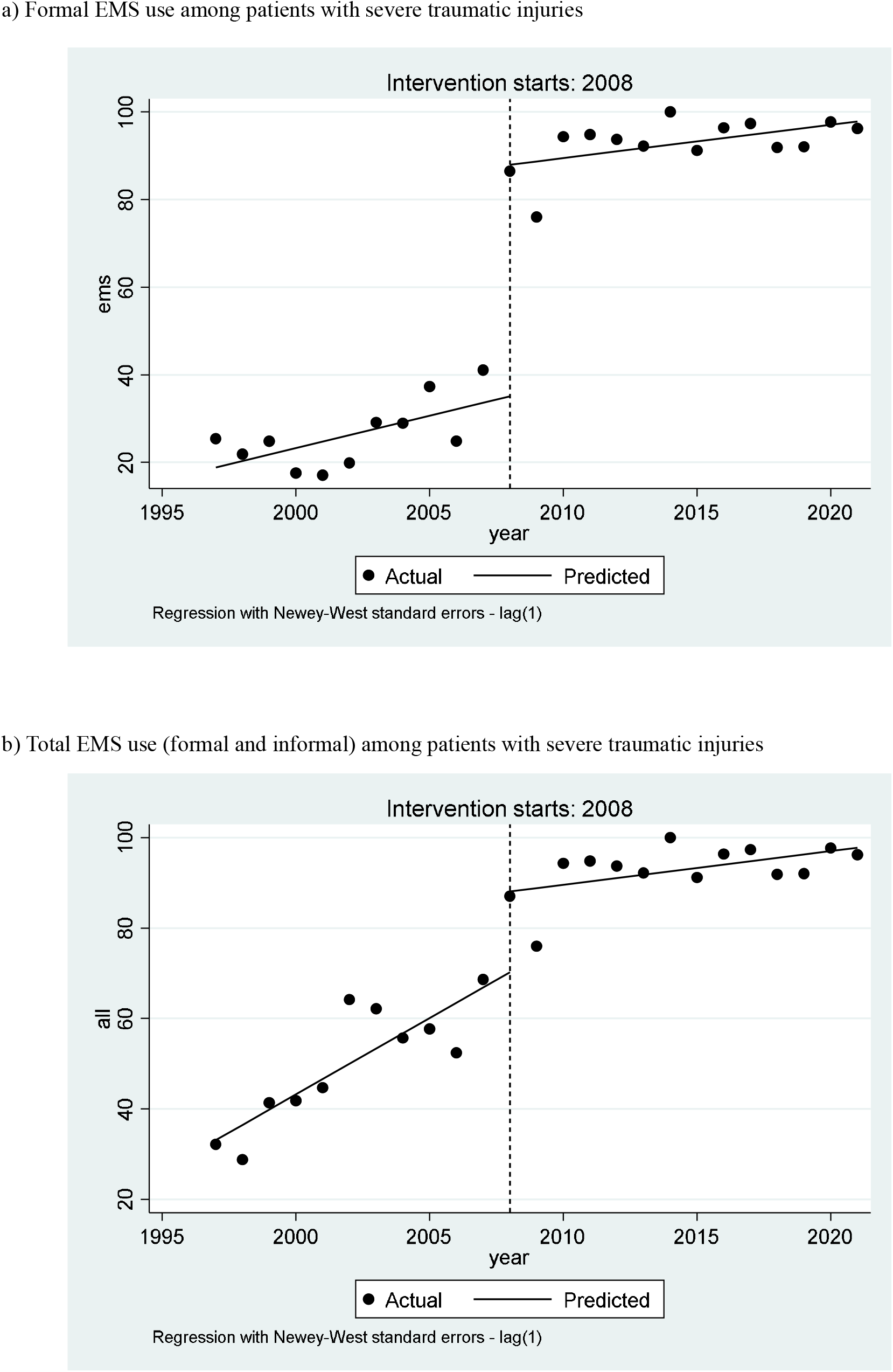
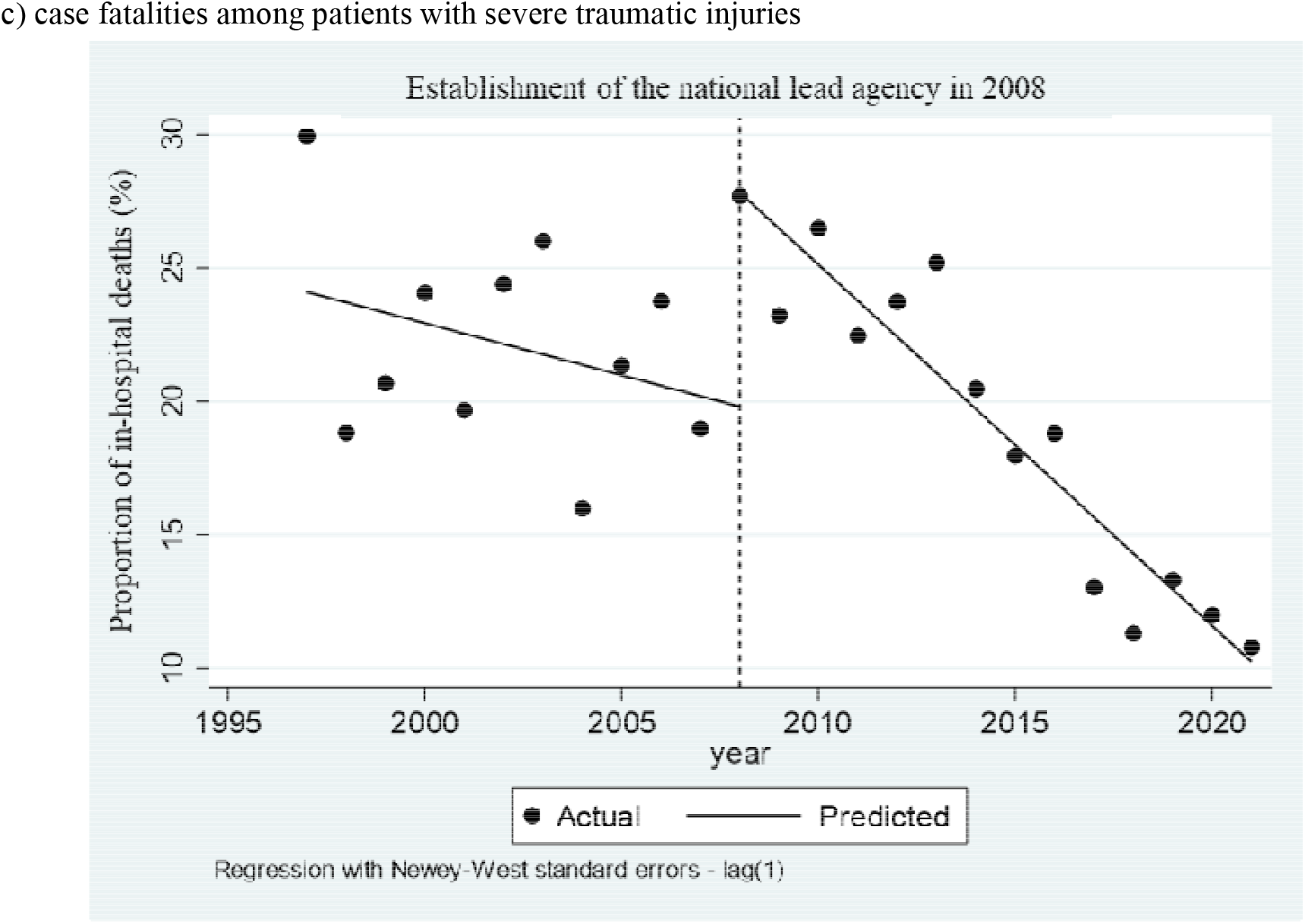
Time trends

The total EMS use, including informal EMS use, increased by approximately 3.4% annually before 2008 (Figure 2b and Table 2). The level change was a significant 17.9 percentage point increase (95% CI=5.9–29.9). A significant slope change was observed in 2008, with a decreasing annual trend.

Before the NIEM establishment, the case fatality rate decreased insignificantly by approximately 0.4% annually; however, there was no downward trend if the data of 1997 was excluded (1997 appeared to be an outlier) (Figure 2c and Table 2). In 2008, there was a significant level change of 8.0 percentage point increase (95% CI= 3.7–12.3). After 2008, the case fatality rate significantly decreased by 1.4% annually (95% CI= -1.6 to -1.1).

## Discussion

In 2008, NIEM was established in Thailand as the national lead agency for the EMS system, according to the Emergency Medical Act. Since then, NIEM has led policymaking, system development, regulation of operations, and coordination with other agencies. While the NIEM develops policies and standards at the national level, local administrators and emergency medical professionals have made significant contributions to the startup, maintenance, and improvement of EMS systems on a provincial basis. For over two decades, Khon Kaen Hospital has greatly contributed not only to the treatment of patients but also to improving the emergency care system, for example, through its contribution towards EMS personnel training.

The present study indicated the impact of the establishment of national lead agency in increasing EMS use among severely injured patients in Khon Kaen though its impact on case fatality was not clear immediately after its establishment. To our knowledge, this is the first study that examined the impact of national lead agency on EMS system using a time series analysis which is more sophisticated in quantifying the effects than simple before-after comparisons.

Although EMS coverage has considerably improved, the increment in formal EMS use among severe trauma patients from 2007 to 2008 was unrealistically large. Achieving such an increase requires creating many new EMS units within a short time, which is not feasible. Informal EMS units, which were numerous at that time, were incorporated into the provincial EMS system as formal units by enrolling them in standardized training, certification, and registration. Before 2008, most patients with severe trauma were transported by informal units run by volunteers. Before the Emergency Medical Act was enacted, emergency patient transport was provided by voluntary/charity organizations in Thailand without formal training, certification, or registration.(7) Integration of the informal sector into the formal system could complement insufficiencies in EMS resources in a short time while standardizing training to ensure quality of care.

A rapid increase in EMS use alone may not be sufficient to mature regional trauma systems to reduce fatalities. According to a systematic review, the fatalities of traumatically injured patients decline depending on the degree of maturity of the trauma system.(18) The starting point of the decline in case fatality rates did not coincide with the rapid increase in EMS use, and seemed to decrease a few years later. It is likely that NIEM’s activities could successfully increase EMS resources in a short time. In contrast, the improvement of regional trauma systems, including pre-hospital and in-hospital trauma care, requires at least several years. For example, improving coordination among different levels of EMS units and introducing fast-track protocols for severe cases after arriving at the emergency department requires time for design and proficiency.

The increase in the case fatality trend observed in 2008 might be an artifact caused by an apparent declining trend resulted from the exceptionally high case fatalities in 1997. Except for this year (1997), the rates were approximately 20%, and 1997 appeared to be an outlier. No abrupt changes were observed in 2008. In the first year (1997) of data collection, non-fatal cases may have been missed from the registry, resulting in a higher fatality rate.

The rapid increase in EMS coverage indicated in the present study might not be replicated in other provinces of Thailand because of the non-uniform patterns of EMS networks by province. The EMS in Khon Kaen Province was the first systematic system to serve as a model for other provinces in Thailand, and other provinces developed their EMS systems following this model.(14, 19) Khon Kaen province has comprehensive management in EMS networks and emergency care system, responding quickly to the policy change.(19) Same or similar increase in EMS use may not have occurred immediately after 2008 in other provinces with less effective systems management capabilities. However, this study’s findings can be generalized to other provinces and countries if the strategies adopted in Khon Kaen after establishing the NIEM in 2008 are followed. This could be an example for other provinces in Thailand and other low- and middle-income countries, which are still developing emergency care systems with limited resources.

Pre-hospital care, which is an important component of the trauma system, may significantly impact trauma mortality, particularly in the indigenous healthcare systems of remote areas, in which transport to the hospital takes hours. Investment in emergency transport systems with basic pre-hospital care alone is promising in these areas. In a ten-year results study in Iraq, trauma mortality was reduced from 17% to 4% by improving pre-hospital trauma care using basic-level personnel (trained volunteers).(20) Sufficient life support measures can improve physiological severity indicators. In Khon Kaen Province, however, a well-organized healthcare system exists, access to hospitals is quite good, and improvement of the entire system, including in-hospital care, is required to reduce fatalities.

## Limitations

This study used secondary data from the Khon Kaen Hospital Injury Surveillance database, which does not register all trauma patients in Khon Kaen Province. However, Khon Kaen Hospital is the only trauma center in the province, and severely injured patients are transported to this hospital. In addition, the role of hospitals in the province did not change over time. Therefore, we can assume that the changes identified in the time-series analysis reflect only policy changes. This study could obtain only 25-year data from 1997 when the surveillance program started. There were no electronic medical records available before 1997. We had at least 10-year data for the pre-policy change period and could identify significant changes after 2008.

## Conclusion

The trends show a significant increase in EMS use among patients with severe trauma after the establishment of the NIEM in 2008, which was attributed to the integration of informal EMS units into the formal system. However, we did not identify a reduction in fatalities in 2008. The increase in EMS use reflects the effect of the national led agency, NIEM, which plays an essential role in shaping national policy and emergency care enhancement.

## Data Availability

All data produced in the present work are contained in the manuscript.

